# Postpartum Psychosis: could genetic vulnerability to insomnia or short sleep duration be protective?

**DOI:** 10.1101/2024.12.13.24319005

**Authors:** Chiara Petrosellini, Sofia H Eriksson, Nicholas Meyer, Olivia Protti, Nicholas Bass, Karoline Kuchenbaecker, Dimitrios Siassakos, Andrew McQuillin

## Abstract

Postpartum Psychosis (PP) is a severe and understudied perinatal mental illness which disproportionately affects women with bipolar disorder (BD). A relationship between sleep disturbance and PP is often assumed, but is poorly understood. From a cohort of 2099 individuals with BD, 343 parous women were identified and screened for perinatal psychiatric complications. We compared 117 women who developed PP with 226 who did not. Polygenic Risk Scores (PRS) for BD, schizophrenia, insomnia, short sleep, long sleep, sleep efficiency and sleep duration were computed using PRS-CS. Logistic regression was used to model the effect of each PRS on PP. Higher PRS for insomnia and short sleep were associated with reduced risk of PP. Individuals in the lowest decile for insomnia PRS (RR 1.96, 95% CI 1.25-3.07, p = 3.50 × 10=:³) and short sleep PRS (RR 2.23, 95% CI 1.40-3.54, p = 7.94 × 10=:=:) had approximately double the risk of PP than individuals in the highest decile. The other PRS were not associated with PP. Mendelian Randomisation analyses did not support a causal relationship between sleep traits and PP. However, we demonstrate that the integration of PRS with bipolar subtype can improve prediction accuracy. Individuals with genetic vulnerability to insomnia or short sleep may develop a heightened tolerance to sleep disruption earlier in life, mitigating the impact of childbirth on mood. These findings suggest that genetic susceptibility to sleep disturbance may be important in the aetiology of PP, offering a new potential avenue for risk stratification and targeted prevention.

## Introduction

Postpartum psychosis (PP) is a severe but treatable mental illness which affects 1-2 in 1000 women who give birth. Most episodes present with mania or a mixed affective state, often accompanied by the presence of psychotic or cognitive symptoms(1). PP is a psychiatric emergency due to its rapid onset and escalation, coupled with the associated risks of suicide and infanticide(2).

Whilst the disease classification of PP is a source of ongoing debate(3), it is widely conceptualised as a form of bipolar disorder (BD) with a vulnerability to childbirth. Women with BD carry a 20% risk of developing a psychotic or manic episode postpartum(4), and a third of women who experience first-onset PP will be diagnosed with BD long-term(5). Despite the close relationship between the two conditions, a previous psychiatric history does not explain most cases of PP and only a proportion of women with BD experience a postnatal relapse. Improved prediction of PP therefore requires better understanding of the role of other perinatal triggers.

Sleep is a fundamental physiological necessity which is almost universally disrupted around childbirth. Pregnancy and the postpartum period are times of extreme change in sleep quantity, quality and pattern(6). There is increasing recognition that these changes can represent both symptoms and causes of maternal mood and anxiety disorders(7). Outside the perinatal period, sleep loss is a well-established trigger of psychosis(8) and a recognised risk factor for suicidality(9).

Experimentally-induced sleep deprivation has been shown to induce mania in women with postnatal psychosis(10) and cause a switch from depression to mania(11). Reduced need or ability to sleep is one of the earliest and most common presentations of PP(12) and sleep has been hypothesized to be the final common pathway through which other factors produce psychosis in susceptible women(13, 14).

There is increasing evidence that sleep may play a role in perinatal episodes for women with BD. Parous women with BD who report sleep loss as a lifetime trigger of manic episodes have been shown to be twice as likely to have had PP as women with BD who have had children and do not report sleep loss as a personal trigger(15). A prospective study has also demonstrated that women with BD who lose one or more nights of sleep over the intrapartum period have a significantly higher risk of developing PP compared to those who do not(16). It is plausible that there is a subset of women with BD who are particularly vulnerable to psychiatric deterioration following sleep disruption, whilst others may be more resilient when sleep patterns are disturbed.

Both BD(17) and sleep traits(18) are thought to have polygenic inheritance, whereby the combined effects of multiple common genetic variants contribute to disease risk. The data from genome-wide association studies (GWAS) of complex traits can be used to derive Polygenic Risk Scores (PRS), which provide an estimate of an individual’s relative susceptibility to disease. Whilst a PRS alone has limited practical application at present, the combination of PRS with other risk factors has been increasingly shown to improve outcome prediction and clinical decision-making in psychiatry(19). A PRS approach can also be used to elucidate disease mechanisms and has provided support to the view that BD and first-onset PP may be biologically distinct(20).

BD demonstrates significant heterogeneity in its pathogenesis, clinical presentation and outcomes(21). At present, prediction of PP in women with BD is imprecise. As a result, resource allocation for pregnant women with pre-existing mental illness is also inefficient. In a direct comparison of women with BD who did and who did not develop PP following childbirth, PRS for BD and schizophrenia were found not to differ significantly between the two groups(20). Our initial objective was to replicate this finding. We then sought to establish whether genetic vulnerability to sleep disorders can be used to improve prediction of PP in women with BD. We chose to investigate the following sleep traits, which we hypothesized would be associated with differing genetic risk profiles in women with BD who develop PP and those who do not:

1. **Insomnia:** difficulty in initiating or maintaining sleep, despite adequate opportunity to sleep(22). Observational(12, 23) and qualitative(24, 25) studies have consistently identified insomnia as one of the most common symptoms of PP. In prospective studies of high-risk cohorts, insomnia has been shown to predict the emergence of BD(26) and has been associated with an increased likelihood of developing delusions and hallucinations(27).
2. **Short Sleep:** commonly defined as habitual sleep duration of less than 6 hours per night(28). Reduced sleep duration is often a prodrome of mania(29) and has been shown to predict deterioration in psychosis symptoms(30).
3. **Long Sleep:** commonly defined as habitual sleep duration of more than 9 hours per night(31). Individuals with BD have been shown to have significantly longer sleep duration than healthy controls, both in periods of euthymia(32) and during depressive relapses(33).
4. **Sleep Efficiency:** the proportion of time in bed which is spent asleep(34). There is consistent evidence that individuals with BD have lower sleep efficiency than healthy controls(32) and a recent meta-analysis has supported an association between reduced sleep efficiency and both early and chronic psychosis(35).
5. **Sleep Duration:** the total amount of sleep obtained per nocturnal sleep episode or across the 24 hour period(36). It is consistently reported as a clinically relevant trait across mood episodes and is linked with both onset and recurrence of BD(37). While short and long sleep have more clearly established associations with affective and psychotic disorders, we also examined sleep duration as a continuous trait given its importance in the interplay between sleep and mood(38–40).

## Methods

### Participants

Individuals with a diagnosis of BD based on the International Classification of Diseases, 10^th^ Revision(41) were recruited from National Health Service (NHS) general practices, hospitals and community mental health teams in England. Participants were also recruited through the Bipolar UK charity. This included individuals with Bipolar I Disorder (BDI), Bipolar II Disorder (BDII) and Schizoaffective Bipolar Disorder (SABD). Ancestrally matched healthy controls were recruited from the same study sites in which BD cases were recruited and from the NHS blood transfusion service(42). All controls were screened for the absence of a lifetime history of psychiatric illness including all major psychotic and affective disorders. All BD cases and healthy controls were of White British descent and had at least three out of four grandparents of the same ancestry. Written informed consent was obtained from all participants. The study received NHS Research Ethics Committee and Health Research Authority approval (MREC/03/11/090).

### Study Measures and Definitions

All participants with BD completed semi-structured interviews using the lifetime version of the Schizophrenia and Affective Disorder Schedule (43) and the Operational Criteria Checklist for Psychotic Illness (44). Female participants who have given birth were screened for a history of perinatal psychiatric complications using the International Cohort Collection for Bipolar Disorder (ICCBD) variables checklist(45). In this cohort, Postpartum Psychosis (PP) was defined as an episode of mania, psychosis or psychotic depression following childbirth.

There is no universal consensus on the timeframe which defines the postnatal period, and there is no widely accepted definition for what qualifies as a ‘perinatal’ episode in women with pre-existing mental illness. Whilst the Diagnostic and Statistical Manual of Mental Disorders (DSM) defines this time period as four weeks(46) and the International Classification of Diseases (ICD) defines this as six weeks following delivery(47), perinatal services in the United Kingdom are designed to care for women until at least a year following childbirth(48). This reflects an increasing recognition that the biopsychosocial changes associated with motherhood have profound personal and public health implications which extend well beyond the early postnatal period(49). In research studies, definitions of PP have varied widely, with some restricting onset to within two weeks of delivery and others including all episodes occurring within the first postpartum year(50). Whilst most episodes of PP emerge within the first two weeks postpartum(51), onset beyond three months is well documented.

Such late-onset presentations have been described in both historical and contemporary literature, and have been linked to physiological transitions such as the resumption of menstruation or the weaning process(52). To reflect the full breadth of clinical vulnerability, we included all episodes occurring within the first twelve months postpartum, consistent with standard definitions of the perinatal period(48). We conducted an additional sensitivity analysis in which we only included individuals who met strict ICD definitions for a perinatal episode. The data were coded in a binary format, to separate parous women with BD into two groups: those who had and those who had not experienced PP following childbirth.

### Discovery datasets for sleep phenotypes

Summary statistics of GWAS were obtained for insomnia(53), short and long sleep(54), sleep efficiency and sleep duration(55) to conduct our analyses.

The insomnia GWAS(53) was conducted in participants recruited to the UK Biobank and 23andMe. Insomnia was defined as self-reported difficulty in falling asleep or staying asleep, which was assessed using a single question: ‘Do you have trouble falling asleep at night or do you wake up in the middle of the night?’. Answers to this question were then dichotomised to create a binary phenotype. A total of 593,724 cases and 1,771,286 controls were included in the GWAS. We used summary statistics from a subset of 386,988 European individuals from the UK Biobank (109,548 cases and 277,440 controls) to generate our PRS.

Austin-Zimmerman et al. performed a multi-ancestry GWAS of self-reported short and long sleep duration in adults recruited to the UK Biobank and the Million Veteran Programme(54). Self-reported sleep duration was measured as a continuous trait and reported in hourly increments for a total of 493,142 individuals. Short sleep was defined as habitual sleep duration ≤5 hours, whilst long sleep was defined as habitual sleep duration ≥10 hours. Normal sleep duration was defined as 7-8 hours per night. Their GWAS meta-analyses compared short versus normal sleep duration and long versus normal sleep duration. We used summary statistics from a subset of 445,966 individuals of European descent to calculate PRS for short sleep (47,054 cases and 382,950 controls) and long sleep (15,962 cases and 382,950 controls).

Jones et al. used accelerometer data from 85,670 UK Biobank participants to perform a GWAS of eight objective sleep traits(55). Sleep efficiency was defined as sleep duration divided by the time period of nocturnal inactivity.

This measure was reported as a percentage ratio in 84,810 individuals of European ancestry. We used summary statistics from this subset to generate PRS for sleep efficiency in our sample. The same summary statistics also included data for accelerometer-derived sleep duration in 85,449 individuals, reported as a continuous measure. We used these to generate PRS for objective sleep duration in our sample as a follow-on analysis, to assess whether these findings differed to those based on binary, self-reported measures of sleep duration reported by Austin-Zimmerman et al(54).

The remaining accelerometer-derived sleep traits reported by Jones et al., were not included in our analyses as current evidence does not strongly support their association with BD or psychosis. While other objective sleep parameters such as M10 timing(56, 57), sleep midpoint(58) and sleep fragmentation(59, 60) may play a role in affective or psychotic disorders, the associations between these traits and illness onset or relapse risk are either limited or inconsistent. They were therefore not included in our hypothesis-driven analyses.

### Genotyping

Participants provided blood or saliva samples for DNA extraction and genome-wide single nucleotide polymorphism (SNP) genotyping was performed. The data was generated at the Broad Institute (Boston, USA) using the Illumina PsychArray or the Illumina Global Screening Array (GSA) genotyping platforms. Strict quality control (QC) was performed separately for each platform using PLINK1.9(61). Imputation was undertaken against the Haplotype Reference Consortium panel using the Sanger Imputation Server(62). Details of the genotyping, QC and imputation methods used have been described elsewhere(63).

### Polygenic Risk Score (PRS) Calculation

Summary statistics for BD(17) and schizophrenia(64) were obtained from the Psychiatric Genetic Consortium. Summary statistics for sleep phenotypes were obtained from the GWAS described above. PRS were computed for BD, schizophrenia and our sleep traits of interest in both subjects with BD and healthy controls. These were generated using the PRS-CS auto method(65), which provides a single score for each individual without the need for p value selection thresholds. PRS-CS requires GWAS summary statistics and a linkage disequilibrium reference panel, and we chose to use the European reference panel from the 1000 Genomes Project(66) for the latter.

For each trait of interest, the raw PRS values were normalised into Z scores centred around a mean of 0 and a standard deviation of 1 in the healthy control subjects, allowing direct comparison across our cohorts (GSA, PsychArray and healthy controls). The normally distributed PRS for the PsychArray and GSA datasets were then merged for downstream analysis.

### Statistical Analysis

Data analysis was conducted in R using version 4.3.2(67). Logistic regression analyses were performed to examine the effect of each PRS on PP following childbirth in women with BD. The genotyping platform used and the first three population principal components were added as covariates in all analyses to account for chip and ancestry confounding. To account for multiple testing, we used the p.adjust function in R to apply the Bonferroni correction method(68) to each PRS analysis. The pROC package in R(69) was used to compute the Receiver Operating Characteristic (ROC) curves and Area Under the Curve (AUC) for significant PRS and for bipolar subtype. For each ROC analysis, 95% confidence intervals for AUCs were obtained using 2000 stratified bootstrap replicates, a standard approach for obtaining stable ROC confidence intervals(70). Differences between AUCs were assessed using DeLong’s test for two correlated ROC curves(71).

For sleep traits in which we observed a significant association between PRS and PP diagnosis, we also added BD PRS and schizophrenia PRS Z scores as covariates in the regression model. In doing so, we attempted to account for possible confounding from differences in relative genetic susceptibility to psychotic illness. Individuals with BDI or SABD are thought to have a higher risk of PP following childbirth than individuals with BDII(72). We therefore also adjusted for bipolar subtype in our regression analyses. To further test the validity of our significant findings, we performed an additional sensitivity analysis in which we only included individuals who developed PP within 6 weeks of childbirth in line with the strict ICD definition of a ‘perinatal episode’ and compared their PRS with individuals who did not develop PP following childbirth.

We performed power calculations using the Additive Variance Explained and Number of Genetic Effects Method of Estimation (AVENGEME) package(73) to estimate whether our analyses were sufficiently powered to detect a significant effect. Of note, AVENGEME is designed to perform power calculations for PRS analyses using older p-value thresholding methods, which limits its application to analyses performed with the newer PRS-CS auto method.

### Mendelian Randomisation (MR) Analyses

For sleep phenotypes where we identified significant associations between PRS and PP, we proceeded with two-sample Mendelian Randomisation (MR) studies to examine whether exposure to specific sleep traits may be causally related to our outcome of interest: PP in women with BD. MR is an epidemiological method which uses genetic variants as instrumental variables to assess causal relationships between potential risk factors and an outcome of interest(74). By using genetic variants as natural experiments, this approach reduces bias from confounding and reverse causation, providing more robust evidence for causal inference. The genetic variants used in MR analyses must satisfy three key assumptions: (i) that they are strongly associated with the exposure of interest, (ii) that they are independent of confounders that could influence the outcome, and (iii) that they only influence the outcome through their effect on the exposure(75). While the genetic association with the exposure can be evaluated, the absence of confounding and exclusion restriction cannot be tested directly.

We extracted genome-wide significant, linkage-disequilibrium independent SNPs from sleep GWAS summary statistics and used these as genetic instruments for insomnia(53) and short sleep(54). We then estimated SNP-outcome associations by using individual-level data from our cohort of parous women with BD. All MR analyses were conducted using the TwoSampleMR package in R(76), which uses the Inverse Variance Weighted (IVW), MR Egger, Weighted Median, Simple Mode and Weighted Mode MR methods to test the relationship between the genetic instruments and outcomes of interest. We conducted heterogeneity tests using both MR Egger and IVW methods, and assessed directional horizontal pleiotropy via the MR Egger intercept.

Our findings are reported in accordance with the Strengthening the reporting of observational studies in epidemiology using MR (STROBE-MR) checklist(77). In line with STROBE-MR guidance(78), we limited MR testing to hypotheses supported by our own findings, to minimise the risk of false-positive findings from exploratory analyses.

## Results

PRS were initially generated for 2099 individuals with BD and 1818 healthy controls in our wider cohort.

343 female participants with BD who had given birth at the point of recruitment were included in our final analyses. Of these individuals, 213 (62.1%) met criteria for BDI, 37 (10.8%) met criteria for BDII and 28 (8.2%) met criteria for SABD. In 65 (19.0%) participants, the BD subtype could not be specified. Of all parous women with BD, 117 had experienced PP following childbirth at the point of recruitment and 226 had not. Those who developed PP had symptom onset between one day and 8 months postpartum. Participants with an underlying diagnosis of a high-risk Bipolar subtype (BPI or SABD) were more likely to develop PP than those without(p=1.387×10^−4^). Women who did and who did not develop PP did not significantly differ in their age or parity at the point of recruitment.

A demographic summary of individuals included in our analyses is shown in Table 1. There was no significant difference in the distribution of genotyping platform used in participants who did and who did not develop PP following childbirth (Chi-squared= 2.717, df= 1, p= 0.099).

**Table 1:** Demographic summary of parous female participants with Bipolar Disorder.

| Demographics | All (n=343) | PP following childbirth (n=117) | No PP following childbirth (n=226) | p value |
| --- | --- | --- | --- | --- |
| Age<sup>1</sup> | 51(16) | 53(14) | 50(16) | 0.684<sup>3</sup> |
| Bipolar Subtype: |  |  |  |  |
| *BPI* | 213 | 86 | 127 | **1.387×10<sup>-4</sup>** |
| *SABD* | 28 | 12 | 16 |  |
| *BPII* | 37 | 3 | 34 |  |
| *BPNOS* | 65 | 16 | 49 |  |
| Parity<sup>2</sup>: |  |  |  |  |
| 1 | 124 | 44 | 80 | 0.776<sup>5</sup> |
| ≥2 | 219 | 73 | 146 |  |

Notes: BPI= Bipolar I Disorder; BPII= Bipolar II Disorder; SABD= Schizoaffective Bipolar Disorder; BP NOS= Bipolar Disorder not otherwise specified
<sup>1</sup> Median age (interquartile range) at recruitment
<sup>2</sup> Defined as the total number of pregnancies carried to a viable gestation at the point of recruitment
Significance set at p < 0.05 (significant findings shown in bold):
<sup>3</sup> Wilcoxon rank sum test comparing age between the two groups
<sup>4</sup> Pearson's Chi-squared test comparing the prevalence of high-risk Bipolar subtypes (BPI or SABD) between the two groups
<sup>5</sup> Pearson's Chi-squared test of independence comparing parity between the two groups

Individuals with lower insomnia PRS were found to be at increased risk of PP than individuals with higher PRS. Each one standard deviation increase in insomnia PRS was associated with an odds ratio of 0.718 for PP (95% CI 0.575-0.897; p=3.5 × 10 ^−3^). This finding remained significant despite stringent correction for multiple testing (Table 2). Each one standard deviation increase in insomnia PRS was therefore associated with an almost 30% decrease in the odds of developing PP. This association remained significant when BD PRS, schizophrenia PRS and bipolar subtype were added as a covariates in the logistic regression model. Women in the bottom decile for insomnia PRS had approximately twice the risk of PP than those in the top decile (RR 1.96, 95% CI 1.25-3.07; Table 3). The distribution of insomnia PRS in women who did and who did not develop PP is shown in Figure 1A.

**Figure 1:**
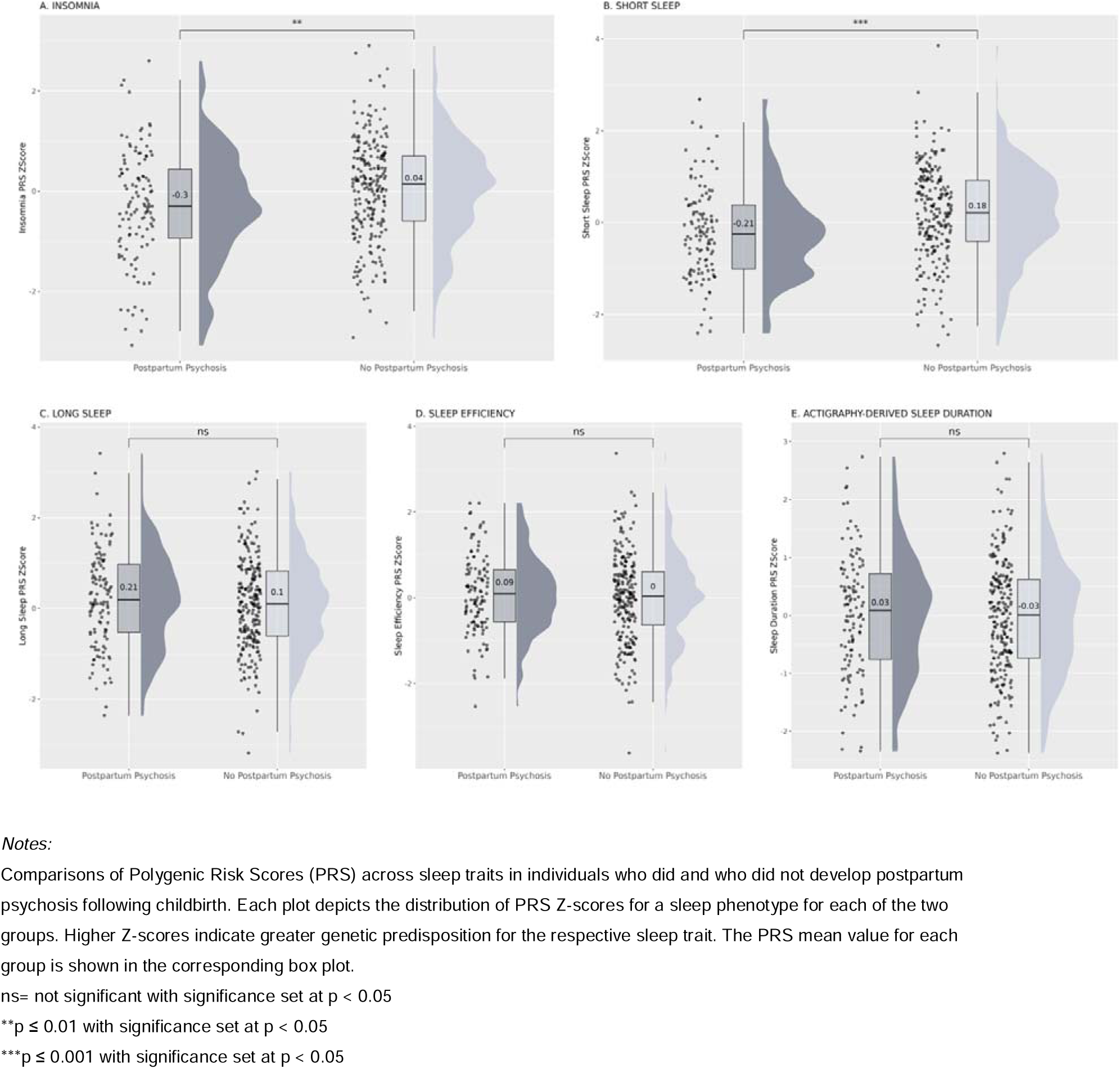
Sleep trait PRS distributions in parous women with bipolar disorder.

**Table 2:** Logistic regressions between PRS and Postpartum Psychosis in parous women with Bipolar Disorder.

| PRS | Odds Ratio (95% CI) | p value<sup>1</sup> | Bonferroni_p<sup>1</sup> |
| --- | --- | --- | --- |
| Bipolar Disorder | 1.255 (0.988-1.594) | 0.063 | 0.441 |
| Schizophrenia | 1.158 (0.922-1.454) | 0.207 | 1.000 |
| Insomnia | 0.718 (0.575-0.897) | **3.50 × 10<sup>-3</sup>** | **2.45 × 10<sup>-2</sup>** |
| Short Sleep | 0.674 (0.535-0.849) | **7.94×10<sup>-4</sup>** | **5.56 × 10<sup>-3</sup>** |
| Long Sleep | 1.100 (0.887-1.365) | 0.384 | 1.000 |
| Sleep Efficiency | 1.074 (0.851-1.356) | 0.546 | 1.000 |
| Sleep Duration | 1.056 (0.846-1.317) | 0.630 | 1.000 |

Notes: Logistic regression analyses performed with Postpartum Psychosis diagnosis following childbirth as the outcome variable and adjusted for 3 population principal components and genotyping platform.
PRS= Polygenic Risk Score; Bonferroni\_p= p value adjusted using the Bonferroni method to correct for multiple testing
<sup>1</sup>Significance set at p < 0.05 (significant findings shown in bold)

**Table 3:** Risk of Postpartum Psychosis according to sleep PRS decile.

| Variable | Risk in lowest decile<br>(%) | Risk in highest decile<br>(%) | Relative Risk<sup>1</sup> (95%<br>CI) |
| --- | --- | --- | --- |
| Insomnia PRS | 27.6 | 14.1 | **1.96 (1.25-3.07)** |
| Short Sleep PRS | 29.3 | 13.1 | **2.23 (1.40-3.54)** |

Notes: Risk of Postpartum Psychosis according to insomnia/ short sleep PRS decile, assuming a background risk of 20% in individuals with bipolar disorder(4, 72)
<sup>1</sup> Relative risk of Postpartum Psychosis in bottom decile compared to top decile for sleep trait PRS (significant findings shown in bold)

We performed a sensitivity analysis comparing individuals who did not develop PP with a subset of individuals who met strict ICD-11 criteria for PP (onset within six weeks postpartum). Whilst this sample size was underpowered to detect a statistically significant effect, the direction of association remained unchanged, with insomnia PRS being lower in individuals who developed PP (mean Z-Score -0.37, SD 0.84) compared to those who did not (mean Z-Score 0.04, SD 1.01).

Individuals with lower short sleep PRS were also found to be at increased risk of PP compared to individuals with higher PRS. Each one standard deviation increase in short sleep PRS was associated with an odds ratio of 0.674 for PP (95% CI 0.535-0.849; p=7.94×10^−4^). This result withstood Bonferroni correction for multiple testing (Table 2). Each one standard deviation increase in short sleep PRS was therefore associated with a decreased odds of PP by approximately one third. As with insomnia, the association between short sleep PRS and PP remained significant when BD

PRS, schizophrenia PRS and bipolar subtype were added as covariates in the logistic regression analysis. Women in the bottom decile for short sleep PRS had over twice the risk of PP than those in the top decile (RR 2.23, 95% CI 1.40-3.54; Table 3). The distribution of short sleep PRS in individuals who did and who did not develop PP following childbirth is shown in Figure 1B. A sensitivity analysis in which only individuals meeting ICD-11 criteria for PP were included as cases was underpowered to detect a statistically significant effect, however the direction of effect was unchanged. Short sleep PRS was found to be lower in individuals who developed PP (mean Z-Score -0.58, SD 0.86) compared to those who did not (mean Z Score 0.17, SD 1.02).

Combining the data for insomnia and short sleep PRS improved prediction of PP in our cohort compared to using bipolar subtype alone, as demonstrated by the increased area under the curve (AUC) in the joint analysis (Figure 2). A model including bipolar subtype alone yielded an AUC of 0.602 (95% CI: 0.555-0.648), whereas adding insomnia and short sleep PRS increased the AUC to 0.697 (95% CI: 0.637-0.753). DeLong’s test confirmed that this improvement in discrimination was statistically significant (Z= –3.91, p = 9.29 × 10=:=:). This suggests improved predictive accuracy when polygenic risk information is included alongside clinical factors.

**Figure 2:**
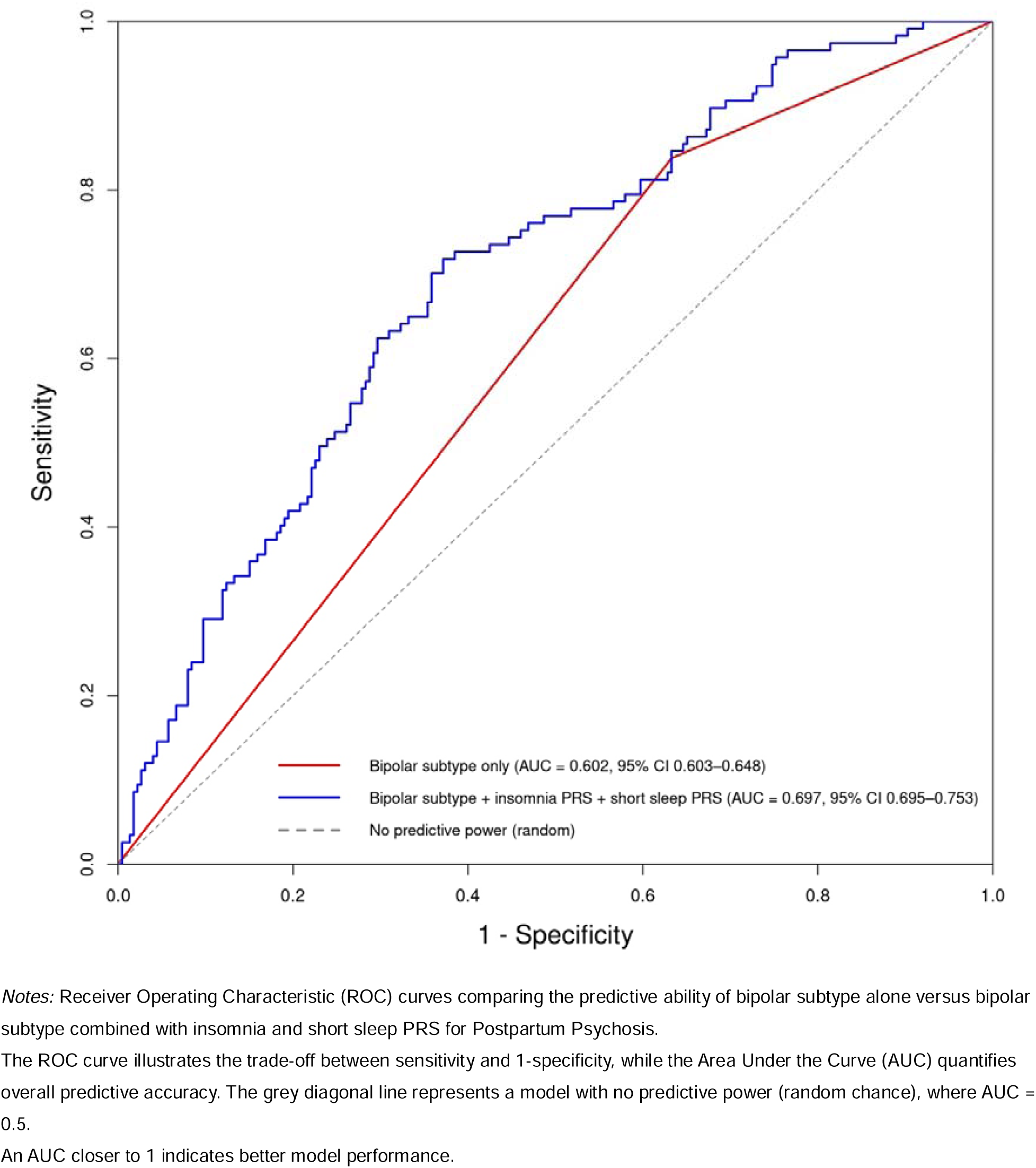
Prediction of Postpartum Psychosis using bipolar subtype alone and in combination with PRS.

Logistic regression comparing individuals with BD who did and who did not develop PP following childbirth did not identify a significant association between BD PRS and PP or between schizophrenia PRS and PP (Table 2). We did not identify significant associations between long sleep PRS and PP (Figure 1C), between sleep efficiency PRS and PP (Figure 1D) or between accelerometer-derived sleep duration PRS and PP (Figure 1E) in our cohort. AVENGEME calculations suggested that all our analyses had >80% power to detect a significant effect (p<0.05), even when stringent thresholds are used in the model. The exception to this was long sleep, where our study may have been underpowered to a detect a significant association (p<0.05) between PRS and PP.

Across all MR methods, we found no evidence to support a causal association between insomnia and PP or short sleep and PP (Table 4). The MR analysis for insomnia showed significant heterogeneity in both MR-Egger (Q = 27.50, df = 9, p = 1.16 × 10=:³) and IVW (Q = 29.61, df = 10, p = 9.92 × 10=:=:) tests, indicating potential variability in the causal estimates of the different genetic variants. We found no evidence to suggest significant horizontal pleiotropy (MR-Egger intercept=0.10, SE = 0.12, p = 0.43).

**Table 4:**
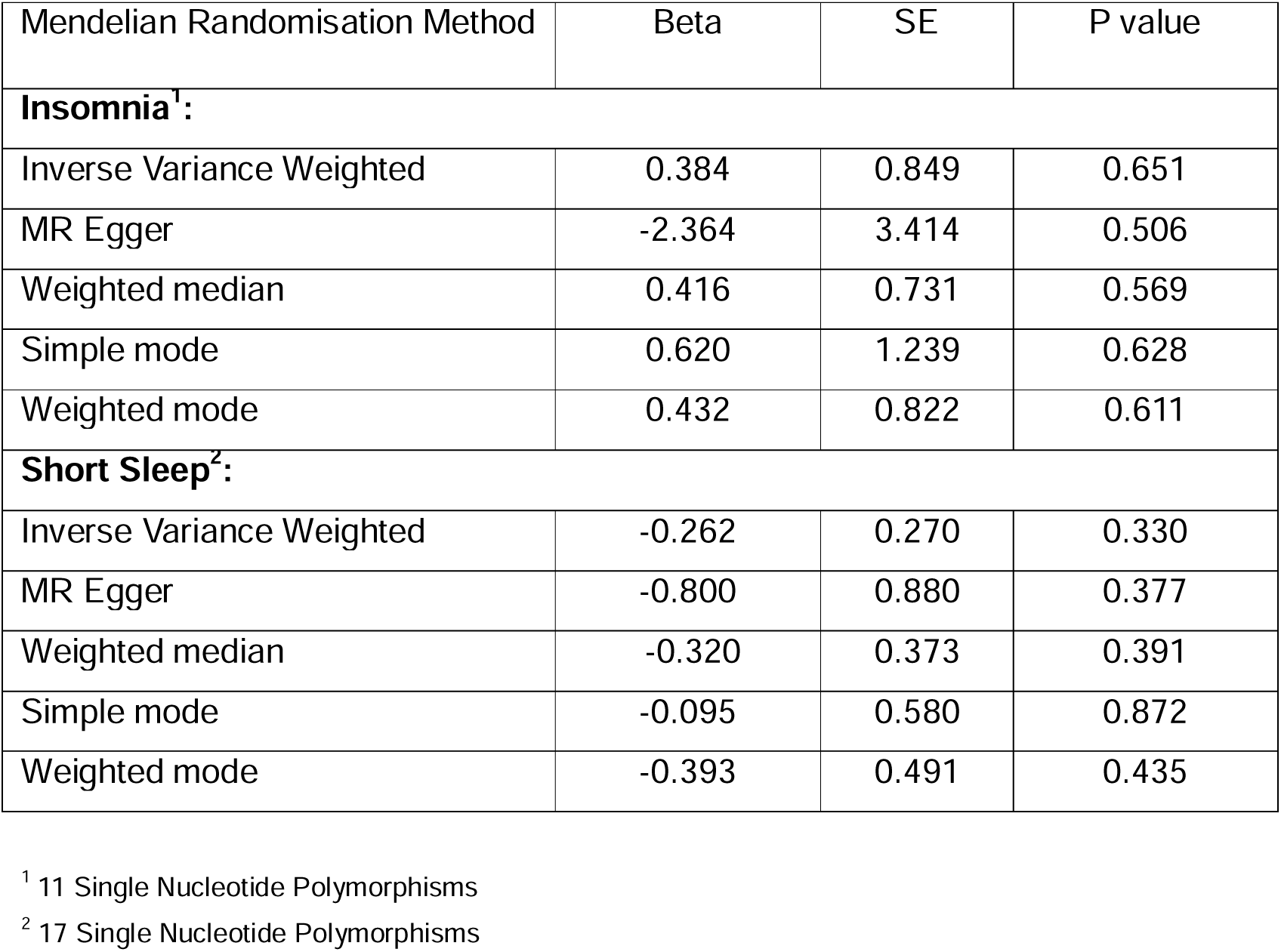
Two-Sample Mendelian Randomisation studies testing causal association between sleep traits and Postpartum Psychosis.

| Mendelian Randomisation Method | Beta | SE | P value |
| --- | --- | --- | --- |
| **Insomnia<sup>1</sup>:** |  |  |  |
| Inverse Variance Weighted | 0.384 | 0.849 | 0.651 |
| MR Egger | -2.364 | 3.414 | 0.506 |
| Weighted median | 0.416 | 0.731 | 0.569 |
| Simple mode | 0.620 | 1.239 | 0.628 |
| Weighted mode | 0.432 | 0.822 | 0.611 |
| **Short Sleep<sup>2</sup>:** |  |  |  |
| Inverse Variance Weighted | -0.262 | 0.270 | 0.330 |
| MR Egger | -0.800 | 0.880 | 0.377 |
| Weighted median | -0.320 | 0.373 | 0.391 |
| Simple mode | -0.095 | 0.580 | 0.872 |
| Weighted mode | -0.393 | 0.491 | 0.435 |

<sup>1</sup> 11 Single Nucleotide Polymorphisms
<sup>2</sup> 17 Single Nucleotide Polymorphisms

For short sleep, we found no evidence of significant heterogeneity using MR-Egger (Q = 5.78, df = 15, p = 0.98) or IVW (Q= 6.19, df = 16, p = 0.99) tests. Additionally, the MR-Egger intercept was 0.03 (SE = 0.05, p = 0.53), indicating no evidence of directional horizontal pleiotropy.

## Discussion

To the best of our knowledge, this study is the first to test the hypothesis that sleep PRS may contribute to perinatal risk stratification in women with BD. We identified that individuals with lower PRS for insomnia and short sleep were at significantly higher risk of developing PP. Women in the bottom decile for each trait had approximately double the risk of PP compared to those in the top decile. Robust statistical corrections were applied to account for potential confounders including bipolar subtype, other PRS, and multiple testing, strengthening the validity of these results. Our findings support previous work indicating that PRS of sleep phenotypes may contribute to risk stratification in BD outside the perinatal period(79). These results suggest that incorporating sleep-related PRS could enhance clinical risk stratification models and inform future preventative approaches.

A genetic correlation (r_g_) of 0.624 has been reported between insomnia and short sleep(54), suggesting that genetic variants associated with one trait are likely to also influence the other to a considerable extent. Notably, the strongest associations in the short sleep GWAS were identified in SNPs in close proximity to *TCF4* and *FOXP2*, both encoding transcription factors which have also been implicated in insomnia(53). If genetic vulnerabilities to self-reported insomnia and short sleep play a role in PP risk, they may therefore do so through shared biological mechanisms. Interestingly, emerging evidence suggests that both *TCF4* and *FOXP2* regulate neurodevelopmental pathways relevant to psychosis. *TCF4* is essential for normal brain cortical development(80) and has been implicated in schizophrenia(81) and major depression(82). *TCF4* was identified as the only gene mediating the bidirectional causal relationship between insomnia and major depression in a MR study(83), and forebrain overexpression of *TCF4* in mice has been shown to increase arousal and shorten sleep duration(84). Similarly, *FOXP2* regulates genes involved in neuronal differentiation, cortical development and synaptic plasticity(85). *FOXP2* polymorphisms have been implicated in dopaminergic signalling, language processing and cognitive function in schizophrenia(86). These findings suggest that genetic influences on arousal, cortical maturation and cognitive regulation may represent a mechanistic framework through which sleep-related vulnerability may influence postpartum psychiatric risk.

Insomnia and short sleep are clinically distinct, with the former representing an internally generated state of hyperarousal leading to subjective perception of poorer sleep quality. This is not reliably associated with objective short sleep duration(87), which can also be due to choice or external circumstances. Nevertheless, the GWAS we used to generate PRS for these phenotypes are both based on self-report, thereby capturing individuals’ subjective experiences of their sleep. Individuals who are biologically more prone to experience insomnia or short sleep may develop a heighted tolerance to sleep disruption over time. It is plausible that the impact of acute postnatal sleep loss on mood stability may be mitigated in women with BD who subjectively experience chronic sleep restriction prior to childbirth. In the pathogenesis of PP, this would suggest that psychiatric deterioration is not determined by sleep loss per se, but rather by the novelty associated with sleep disruption. This interpretation is consistent with the observation that primiparity is a known risk factor for PP(88), as first-time mothers may experience a heighted response to sleep loss compared to multiparous women who would have previously been exposed to perinatal sleep disruption.

Perceived adjustment to chronic sleep loss is often a misperception, and subjective adaptation may occur despite cumulative physiological impairment and sustained sleep pressure(89–91). Nevertheless, this psychological adaptation may play a role in affective resilience. There is growing interest in the concept of sleep resilience, which describes the capacity to maintain emotional, cognitive and physical functioning when sleep or circadian patterns are disturbed(92). Significant individual differences in vulnerability are observed and resilience in one functional domain does not necessarily extend to others(93). While resilience is known to moderate the cognitive consequences of poor sleep(94, 95) and mediate the relationship between sleep and mental illness(96–98), it is uncertain whether this adaptability can be acquired through chronic sleep disturbance. Vulnerability to sleep loss has been described as trait-like, as individual behavioural responses appear to be relatively stable across both acute and more prolonged sleep restriction(99–101). However, the existing evidence comes from small studies with brief observation periods which do not capture the cumulative impact of lifelong sleep behaviours or inherited vulnerability. A computational modelling study simulated the long-term effects of habitual sleep patterns on responses to acute sleep deprivation, identifying that short sleepers were more resilient and had longer duration of waking episodes following recovery sleep(102). This is consistent with evidence that habitual short sleepers may be more tolerant of homeostatic sleep pressure(103). While there is no longitudinal evidence that genetic vulnerability to insomnia or short sleep mitigates the effects of sleep loss on mood, our findings raise the possibility that these traits may confer resilience in the context of PP.

Our analyses suggest that incorporating PRS for insomnia and short sleep alongside clinical variables may improve the accuracy of PP risk prediction. Bipolar subtype(72, 104), use of prophylactic medication(4, 105), perinatal psychiatric history(72), and family history(106, 107) are important clinical predictors of PP. Whilst data was not available for all of these factors in our cohort, future predictive models should aim to integrate these clinical variables with PRS information. This combined approach could improve risk stratification and represents a promising step toward more personalised preventive strategies for women at high risk of PP. At present, however, PRS are not yet suitable for clinical implementation due to their well-recognised limitations(108). Therefore, the translational implication of this work lies in advancing mechanistic understanding and generating hypotheses, rather than suggesting imminent clinical adoption.

Our MR analyses do not support a causal relationship between insomnia and PP or short sleep and PP, indicating that the observed associations may reflect correlated genetic or environmental vulnerabilities rather than direct causation. However, these null findings may also have been influenced by the weak genetic instruments available and the heterogeneity among them. The relatively low estimated SNP heritability of our traits of interest (8.2% for insomnia(53) and 11.9% for short sleep(54)) also somewhat violates the first assumption of MR. Furthermore, it is important to note that these sleep traits are highly polygenic, and PRS will therefore capture genetic susceptibility much more accurately than an MR model with relatively few variants. Whilst pre-existing insomnia or short sleep may represent a useful clinical marker in perinatal risk assessment, there is insufficient evidence to support a causal association between genetic predisposition to these traits and PP at present.

We did not find evidence to suggest a significant association between long sleep PRS and PP risk. Women who experienced PP had higher PRS for long sleep than participants who did not, albeit below the threshold for statistical significance. This is in keeping with the hypothesis that individuals who are chronically exposed to sleep restriction prior to childbirth may be less vulnerable to PP. Conversely, women who are biologically predisposed to habitual long sleep may be more affected by acute perinatal sleep loss. Our study lacked sufficient power to test this theory, in part because long sleep has a lower SNP heritability compared to the other sleep phenotypes examined(54), resulting in reduced predictive power of its PRS. Improved GWAS of long sleep may offer the opportunity to robustly test this notion in future studies.

Our study did not identify an association between sleep efficiency PRS and PP outcome. Reduced sleep efficiency is often observed in actigraphy studies of individuals with BD(32) and has been implicated in psychosis relapse(35). Recent evidence also suggests that higher sleep efficiency at baseline predicts lower subjective sleepiness during prolonged sleep deprivation, indicating a potential role in mood regulation(109). It is plausible that women who are able to effectively capitalise on available sleep opportunities may experience less mood disturbance from perinatal sleep disruption, whilst those with lower sleep efficiency may be more susceptible to psychiatric deterioration. Study designs which integrate subjective and objective assessments of sleep efficiency may be helpful to test this theory further in perinatal cohorts.

While we found a significant association between PRS for self-reported short sleep and PP, we did not identify a significant association between accelerometer-derived sleep duration PRS and PP. This may in part reflect the weak genetic correlation between self-reported and objectively measured sleep duration observed in the discovery GWAS (r_g_=0.43) (55). Observational data also show that objective and subjective sleep measures can differ substantially in the same individual, with studies frequently reporting low concordance between self-reported and actigraphy-derived estimates of sleep duration(110, 111). In perinatal populations, subjective perceptions of sleep may be more closely linked to mental health outcomes than objective measures. Several studies have found self-reported sleep measures to be more predictive of postpartum depression than any actigraphy-derived metric(112–115). Taken together with our results, these findings suggest that subjective perceptions of sleep (and the genetic liability underlying them) may be more clinically relevant than objective sleep measures in predicting vulnerability to perinatal mood disorders.

Despite our study having adequate power to detect a significant association, we found no significant difference in BD PRS or schizophrenia PRS in women who did and who did not develop PP following childbirth. Our results are consistent with previous work in which BD and schizophrenia PRS were compared in parous women with BD who did and who did not develop PP(20). Psychotic symptoms are highly prevalent in BD(116) and women with BD have a significantly increased risk of affective psychoses in the perinatal period(117). The estimated heritability of BD lies in the region of 60-85% and common genome-wide variants are thought to account for a significant proportion of this(17).

Given the substantial genetic contribution to phenotypic variance in BD and the high genetic correlation between BD and schizophrenia(118), it would be plausible that PP may represent a more severe BD phenotype and that PRS associated with psychotic illnesses could be used to improve outcome prediction. Our negative finding may partly reflect the limited explanatory power of current PRS for BD and schizophrenia, or it may indicate that the hypothesis that PP represents a more severe bipolar phenotype is incorrect. Irrespective of the underlying cause, our results and existing evidence indicate that PP cannot simply be explained by individual genetic susceptibility to psychotic illness.

Our findings should be considered within the context of some important limitations. The retrospective cross-sectional design of our study renders it susceptible to recall bias. Data collection primarily relied on participant reports and clinical details could not always be validated through case notes. Additionally, the interviews captured case-control status at a specific point in time, potentially missing PP in subsequent pregnancies among individuals labelled as controls. Despite our evidence-based rationale for including cases occurring up to 12 months postpartum, this broader definition inevitably increases heterogeneity within the PP group. However, this is not a limitation unique to our methodology, as PP is increasingly recognised as a highly heterogeneous clinical entity(119), and the absence of standardised diagnostic criteria continues to hinder research in this area.

While our psychiatric phenotyping was comprehensive, the lack of detailed sleep data limits the conclusions that can be drawn on the relationship between PRS, sleep phenotype and PP outcome. Moreover, sleep-related PRS are relatively weak genetic instruments, and the extent to which they reflect sleep behaviour, particularly in individuals with BD, remains to be clarified. In addition, all available data were based on lifetime, retrospective measures that do not provide sufficient granularity to evaluate potential perinatal-specific differences. Important clinical variables such as medication use and treatment response were recorded without reference to timing in relation to pregnancy or childbirth. The absence of longitudinal data limited our ability to assess whether specific exposures during the perinatal period influenced risk of PP. This represents a limitation of our dataset and highlights the need for prospective studies that capture clinical information specific to the perinatal period. Several important potential confounding factors were also not available in our dataset, including socioeconomic status, smoking, substance misuse and physical health comorbidities, which are known to influence both sleep and mental health and could contribute to the observed associations(120). Future studies incorporating richer clinical and sociodemographic data will be essential to better disentangle these relationships.

Finally, our sample characteristics may also limit the generalisability of our findings. Women with high-risk bipolar subtypes (BPI or SABD) were overrepresented in our study cohort compared to their prevalence the general population(121) and our findings may not be epidemiologically representative of all women with BD. It is important to note that our findings pertain specifically to women with BD and should not be used to draw conclusions on the relationship between sleep PRS and PP risk in low-risk individuals. Furthermore, the exclusive inclusion of individuals of European ancestry restricts the generalisability of findings to other populations. Future work should investigate the role of sleep PRS in women experiencing new-onset PP without a pre-existing history of mental illness to elucidate this relationship further, and should include more diverse populations to improve generalisability.

PP is a complex, multifactorial illness with potentially devastating consequences. Improved prediction of PP is essential to facilitate early intervention and mitigate its impact on maternal, child and public health. In women with BD, prediction of perinatal episodes is particularly challenging due to clinical heterogeneity and controversies in disease classification. We demonstrate that individuals with BD who develop PP may have a lower genetic propensity for insomnia and short sleep. These findings suggest that genetic vulnerability to sleep disturbance may contribute to the aetiology of PP, highlighting the potential value of assessing individual sleep history in perinatal psychiatric risk. In particular, our results prompt further inquiry into whether individuals with a genetic predisposition to chronically short or restricted sleep may be more resilient to the acute sleep loss associated with childbirth. In view of the limitations described above, however, our findings should be considered exploratory and will require replication in larger, well-characterised perinatal cohorts. Future work should prioritise multi-ancestry recruitment, unified definitions of PP and the use of both objective and subjective sleep measures. Prospective studies in women with and without pre-existing mental illness are essential to further understand the role of PRS in perinatal psychiatric risk. These efforts will not only contribute to early intervention and improved management of conditions like PP, but will also drive the ongoing progress of precision medicine in mental healthcare.

## Data Availability

Data are available upon request to the authors

## Acknowledgments

The authors would like to gratefully acknowledge all DNA Polymorphisms In Mental illness (DPIM) study participants for their contribution to this work. Participant recruitment was funded by the Medical Research Council (MRC grant code G1000708), the Neuroscience Research Charitable Trust, the Camden and Islington NHS Foundation Trust, a research lectureship from the Priory Hospitals and the National Institute for Health and Care Research (NIHR) Mental Health Research Network (MHRN). The study also received support from Bipolar UK (formerly the UK Manic Depression Fellowship). Genotyping was funded by the Stanley Center for Psychiatric Research at the Broad Institute. Parts of the analyses were supported by a project that has received funding from the European Research Council (ERC) under the European Union’s Horizon 2020 research and innovation program (Grant agreement No. 948561). CP is supported by the Rosetrees Trust and the Elizabeth Garrett Anderson Hospital Charity. DS is supported by the NIHR. AM, NB and SE are supported by the NIHR University College London Hospitals Biomedical Research Centre funding scheme.

## Data availability

The GWAS summary statistics used for these analyses is publicly available through the original publications for the different phenotypes of interest: bipolar disorder(17), schizophrenia(64), insomnia(53), short and long sleep(54), sleep efficiency and sleep duration(55). The participant data set used for this study as well as the code for the analyses conducted is available upon request.

